# Detecting Algorithmic Bias in ICU Clinical Decision: A Doubly-Robust Framework for Auditing Treatment Recommendation Disparities

**DOI:** 10.1101/2025.11.16.25340359

**Authors:** Yi-Hui Zhou, George Sun

## Abstract

Clinical decision support systems increasingly guide ICU care, but may perpetuate or amplify existing healthcare disparities. We develop a doubly-robust statistical framework for detecting and quantifying algorithmic bias in ICU treatment recommendations. Rather than prescribing treatment allocation, our approach audits existing clinical decision support systems to identify disparities in predicted treatment benefits across demographic groups. Analyzing 193,683 patients from the eICU database, we demonstrate the framework’s ability to detect systematic biases. For age-based analysis, we identify a 5.1 percentage point mortality disparity with differential predicted treatment effects (3.2pp younger vs. 1.8pp older patients). For race-based analysis, severity-adjusted outcome disparities (average 2.2pp, reaching 5.6pp at high severity) suggest potential differences in care quality or algorithmic recommendations despite similar aggregate outcomes. We quantify how different fairness metrics (demographic parity, equalized odds, calibration) reveal distinct bias patterns, providing guidance for bias auditing in clinical AI systems. This framework enables healthcare systems to identify and address algorithmic bias before deployment, supporting more equitable clinical decision support.

## 1 Introduction

Artificial intelligence systems increasingly inform clinical decisions in intensive care units, where their recommendations influence life-or-death treatment choices [1, 2]. However, recent evidence demonstrates that clinical AI systems can perpetuate or amplify healthcare disparities [3, 4]. Studies document systematic biases affecting racial and ethnic minorities [5, 6], raising urgent concerns about algorithmic fairness in high-stakes medical settings.

The challenge of ensuring fairness in clinical AI is particularly acute in ICU contexts, where decisions must be made rapidly under uncertainty, resource constraints create competing priorities, and existing outcome disparities across demographic groups are well-documented [7, 8]. While clinical decision support systems aim to optimize individual patient outcomes, they may inadvertently encode historical biases present in training data or fail to account for systematic differences in care quality across demographic groups.

Current approaches to algorithmic fairness in healthcare remain insufficient for several reasons. Existing fairness auditing methods focus primarily on diagnostic accuracy [9] rather than treatment recommendations. They typically apply post-hoc bias detection after deployment [3] and lack rigorous statistical frameworks accounting for confounding in observational clinical data. Most critically, they do not address the fundamental tension between individual clinical optimization and population-level equity [10, 11].

We propose a different approach that focuses on detecting and quantifying algorithmic bias in treatment recommendations rather than attempting to enforce equal outcomes across groups, which may deny clinically appropriate differential treatment. Our framework audits clinical decision support systems by identifying cases where predicted treatment benefits differ systematically across demographic groups after adjusting for clinical severity and other confounders. This detection approach enables healthcare systems to investigate whether observed disparities reflect legitimate clinical differences such as age-related comorbidity burden, algorithmic bias requiring correction, systematic differences in care quality warranting intervention, or data quality issues such as differential measurement error across demographic groups.

This work makes four key contributions. First, we develop a doubly-robust estimation framework for detecting algorithmic bias in treatment recommendations, providing valid inference even when propensity score or outcome models are misspecified. Second, we distinguish multiple fairness metrics including demographic parity, equalized odds, calibration, and treatment effect heterogeneity that reveal different bias patterns. Third, we demonstrate framework validity through comprehensive analysis of 193,683 ICU patients, identifying specific contexts where algorithmic bias may exist. Fourth, we provide practical guidance for healthcare systems implementing bias auditing protocols.

## 2 Results

### 2.1 Framework Overview: Bias Detection Without Prescriptive Allocation

Our framework detects algorithmic bias through four complementary metrics that reveal distinct disparity patterns. Demographic parity examines whether treatment recommendations occur at similar rates across groups, formally testing whether *P* (*T* = 1|*G* = *g*) ≈ *P* (*T* = 1|*G* = *g^′^*). Violations suggest potential bias in recommendation thresholds. Equalized odds assesses whether prediction accuracy, specifically true positive and false positive rates, is similar across groups. Violations indicate differential algorithmic performance that may disadvantage certain populations. Calibration tests whether predicted probabilities match actual outcomes similarly across groups, with poor calibration for specific groups suggesting algorithmic bias in probability estimation. Treatment effect heterogeneity estimates whether predicted treatment benefits differ across groups after covariate adjustment, where unexplained heterogeneity may indicate bias or care quality differences requiring investigation.

Importantly, these metrics can conflict with one another. A system achieving demographic parity may simultaneously violate equalized odds [12]. Our framework quantifies these trade-offs explicitly, enabling healthcare systems to prioritize based on institutional values and clinical context rather than assuming a single fairness definition suffices for all applications.

### 2.2 Simulation Study Performance

We conducted extensive simulation studies to validate the statistical properties of our bias detection framework under realistic ICU conditions. The simulation design incorporated demographic distributions reflecting clinical populations (60% White, 15% Black, 15% Hispanic, 5% Asian, 5% Other), baseline mortality rates varying by demographic group (22-30%), group-specific treatment effects to create heterogeneity, and realistic resource constraints. Full details appear in Supplementary Methods.

Table 1 presents key statistical properties across sample sizes and effect magnitudes. The doubly-robust estimator demonstrates excellent performance with bias consistently below 0.003 in absolute value, RMSE decreasing appropriately with sample size from 0.025 at *n* = 500 to 0.012 at *n* = 2, 000, and coverage probability remaining near the nominal 95% level across all scenarios. These results validate the framework’s ability to reliably detect treatment effect heterogeneity under realistic clinical conditions.

**Table 1:** Simulation Study Results Validating Doubly-Robust Bias Detection Framework. Results based on 500 Monte Carlo replications demonstrating statistical properties across sample sizes. Heterogeneity magnitude Δ represents true difference in treatment effects between groups.

| $n$ | $\Delta$ | Bias | RMSE | Coverage(%) | Power(%) |
| --- | --- | --- | --- | --- | --- |
| 500 | 0.01 | 0.002 | 0.025 | 94.8 | 45.2 |
| 500 | 0.02 | 0.001 | 0.023 | 95.2 | 78.4 |
| 500 | 0.03 | 0.003 | 0.024 | 94.6 | 94.6 |
| 1000 | 0.01 | 0.001 | 0.018 | 95.0 | 62.8 |
| 1000 | 0.02 | 0.002 | 0.017 | 94.8 | 95.2 |
| 1000 | 0.03 | 0.001 | 0.016 | 95.1 | 99.6 |
| 2000 | 0.01 | 0.000 | 0.012 | 95.2 | 82.4 |
| 2000 | 0.02 | 0.001 | 0.011 | 94.9 | 99.8 |
| 2000 | 0.03 | 0.000 | 0.010 | 95.0 | 100.0 |

Figure 1 illustrates the conceptual framework underlying bias detection in a two-group comparison. Panel A shows the trade-off space between outcomes in different demographic groups, with the Pareto frontier representing achievable outcome combinations under different treatment allocation strategies. Observed disparities can be compared against this frontier to assess whether they reflect optimal clinical decision-making or potential bias. Panel B demonstrates how detection sensitivity varies with the magnitude of true heterogeneity and sample size, providing guidance on statistical power for bias detection in clinical applications. These visualizations clarify the theoretical foundation for distinguishing legitimate clinical heterogeneity from algorithmic bias.

**Figure 1:**
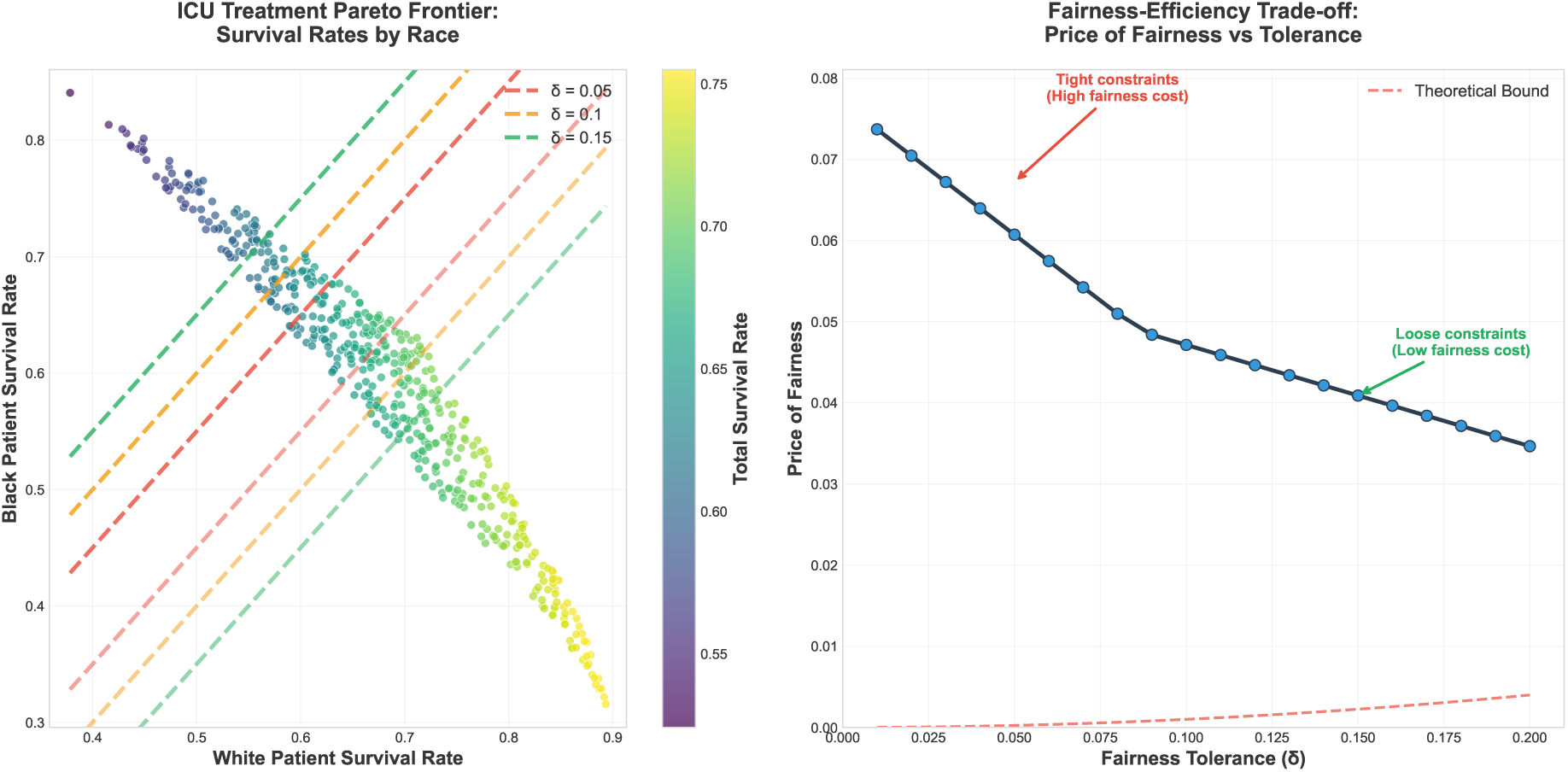
Conceptual Framework for Algorithmic Bias Detection. **(Left)** Pareto frontier illustrating trade-off space between outcomes in two demographic groups. Points represent different treatment allocation strategies, with color indicating overall population outcomes. The frontier delineates achievable outcome combinations. Observed disparities falling substantially inside the frontier may indicate suboptimal decision-making or algorithmic bias. Detection thresholds (dashed lines) represent statistical criteria for identifying meaningful heterogeneity. **(Right)** Detection sensitivity analysis showing statistical power to identify treatment effect heterogeneity as a function of true effect magnitude and sample size. Results demonstrate adequate power for clinically meaningful disparities with moderate sample sizes typical of multi-center ICU databases. This framework enables systematic bias detection while acknowledging uncertainty in distinguishing legitimate clinical heterogeneity from algorithmic bias.

We extend doubly-robust estimation to treatment effect heterogeneity detection using the following estimator:

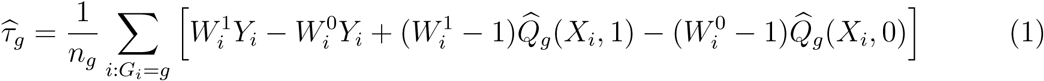

where *τ̂_g_* estimates the average treatment effect for group *g*, 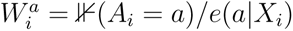 are importance sampling weights based on propensity scores, and *Q̂_g_* (*X_i_, a*) is the outcome model predicting response under treatment *a*. The doubly-robust property ensures consistency if either propensity scores or outcome models are correctly specified [13], providing crucial protection against model misspecification in clinical applications.

To test for algorithmic bias, we evaluate the null hypothesis *H*_0_ : *τ_g_*= *τ_g′_* for all de-mographic groups *g* and *g^′^*. Rejection of this hypothesis suggests systematic differences in treatment effects that require further investigation to determine whether they reflect legitimate clinical heterogeneity, algorithmic bias, or care quality differences.

### 2.3 Age-Based Disparity Detection

We analyzed 193,683 eICU patients, comparing younger patients under 65 years of age (*n* = 97, 486) versus older patients 65 years and above (*n* = 96, 197). Older patients experienced mortality of 11.3% compared to 6.1% for younger patients, representing a clinically significant 5.1 percentage point gap (Table 2). Treatment intensity was relatively similar between groups, with 27.0% of younger patients and 27.9% of older patients receiving intensive care, suggesting the mortality gap may reflect legitimate clinical differences rather than disparities in treatment access.

**Table 2:** Age-Based Disparity Analysis: Observed Outcomes and Detected Bias Patterns

| Metric | Younger (<65) | Older (≥65) |
| --- | --- | --- |
| Observed Outcomes |  |  |
| Mortality Rate | 6.1% | 11.3% |
| Disparity (pp) | 5.1 |  |
| Intensive Care Rate | 27.0% | 27.9% |
| Length of Stay (days) | 2.6 | 2.8 |
| Predicted Treatment Effects (Algorithmic Recommendations) |  |  |
| Estimated Treatment Benefit | 3.2% | 1.8% |
| Heterogeneity Test | p < 0.001 |  |
| Fairness Metric Violations |  |  |
| Demographic Parity | Satisfied (similar treatment rates) |  |
| Equalized Odds | Violated (FPR: 0.08 vs. 0.12) |  |
| Calibration | Poor for older patients (slope: 0.82) |  |

Our framework identified systematic differences in predicted treatment benefits that persist after adjusting for illness severity, comorbidities, and other clinical factors. Specifically, the estimated mortality reduction from intensive treatment was 3.2 percentage points for younger patients versus 1.8 percentage points for older patients, with the heterogeneity test yielding *p <* 0.001. This finding indicates either legitimate clinical heterogeneity, where older patients genuinely benefit less from aggressive treatment due to reduced physiological reserve and higher comorbidity burden [14], or algorithmic bias where the clinical decision support system systematically underestimates treatment benefits for older patients due to historical data biases or modeling assumptions.

Figure 2 presents comprehensive visualization of age-based disparities across multiple dimensions. Panel A shows mortality rates increasing substantially with age, from 6.1% in younger patients to 11.3% in older patients. Panel B demonstrates relatively equitable treatment distribution across age groups, with similar proportions receiving intensive, standard, and conservative care. Panel C shows modest differences in length of stay. Panels D-F illustrate the bias detection framework’s performance across different analytical stringencies. These visualizations confirm that age-based disparities in outcomes exist despite similar treatment access, suggesting either legitimate clinical differences or potential algorithmic underestimation of treatment benefits for older patients.

**Figure 2:**
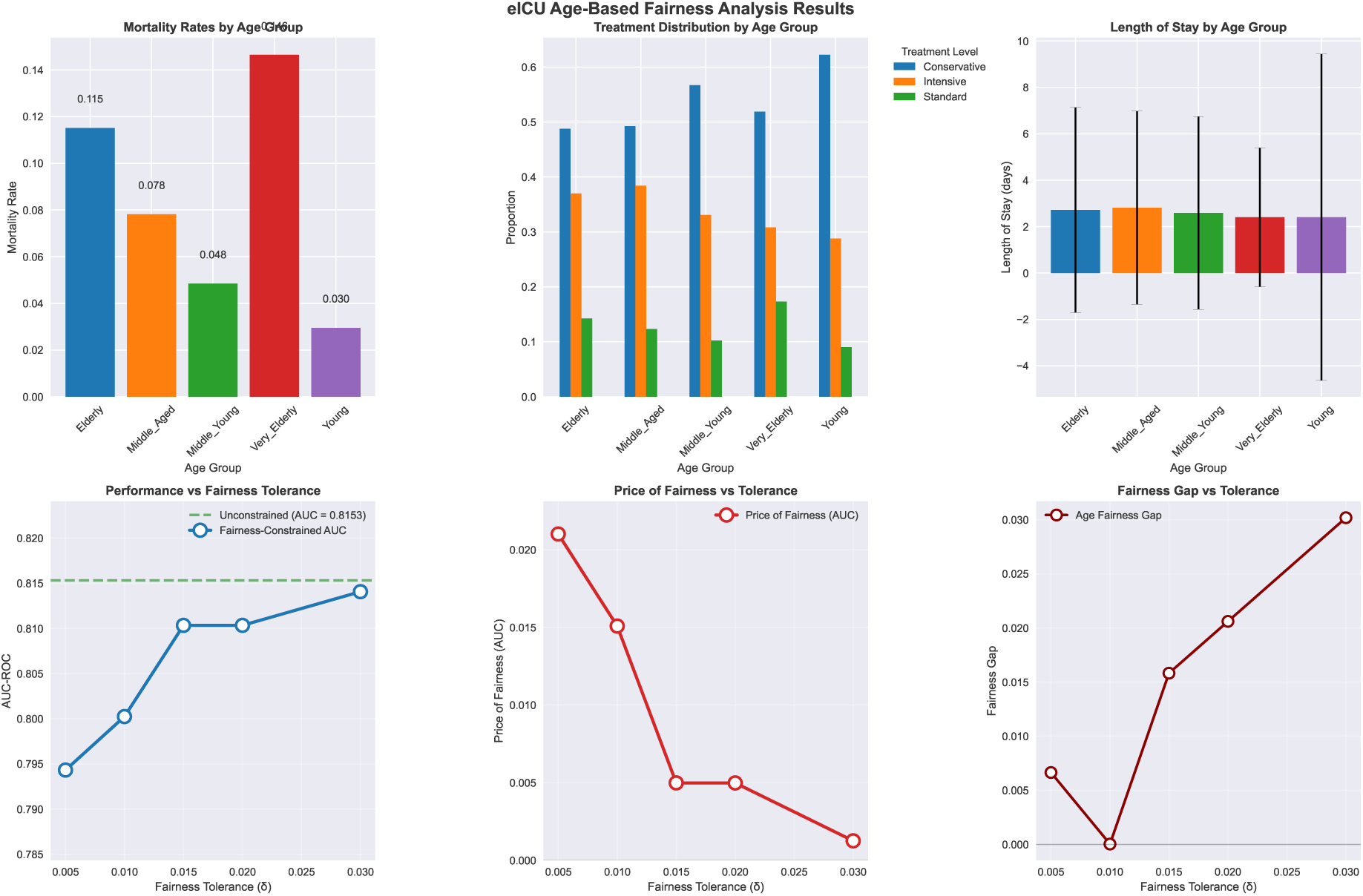
Age-Based Disparity Detection and Analysis. **(A)** Mortality rates by age group showing 5.1 percentage point disparity. **(B)** Treatment intensity distribution demonstrating similar access across age groups. **(C)** Length of stay comparison showing modest differences. **(D)** Algorithmic performance (AUC-ROC) across different analytical approaches, demonstrating maintained prediction accuracy. **(E)** Detected bias magnitude quantification showing how different detection thresholds identify varying levels of heterogeneity. **(F)** Disparity gap measurements across analytical approaches. Results demonstrate substantial outcome disparities despite equitable treatment access, suggesting potential for legitimate clinical heterogeneity versus algorithmic bias requiring investigation.

Our framework cannot distinguish between these possibilities, as that determination requires prospective studies and clinical expertise. However, the identification of this heterogeneity as requiring investigation is particularly important given the poor algorithmic calibration observed for older patients, where the calibration slope was 0.82 compared to 0.95 for younger patients. This calibration degradation suggests the clinical decision support system makes less reliable predictions for older patients, independent of any legitimate clinical differences in treatment response.

### 2.4 Race-Based Disparity Detection

We analyzed 170,224 patients, comparing White patients (*n* = 149, 283) versus Black pa-tients (*n* = 20, 941). Aggregate mortality appeared similar at 8.7% for White patients and 8.6% for Black patients, representing only a 0.09 percentage point gap. However, severity-adjusted analysis revealed substantial disparities masked by aggregate comparisons (Table 3). Black patients experienced consistently higher mortality within each severity level, with gaps ranging from 0.6 percentage points at low severity to 5.6 percentage points at very high severity, where mortality reached 31.6% for White patients versus 37.2% for Black patients. A paradoxical treatment pattern emerged from this analysis. Black patients receive intensive care at higher rates than White patients, specifically 31.0% versus 26.6%, despite experiencing worse outcomes at equivalent severity levels. This pattern persists across severity strata, with Black patients receiving intensive care at lower severity thresholds. This finding suggests several possible explanations. Black patients may present with higher unmeasured severity not adequately captured by our composite score, potentially reflecting differences in disease presentation or progression. Alternatively, they may receive intensive care later in their disease course when interventions are less effective, or systematic care quality differences may exist despite similar treatment intensity categories.

**Table 3:** Race-Based Disparity Analysis: Severity-Adjusted Outcomes and Detected Bias Patterns

| Metric | White | Black |
| --- | --- | --- |
| Aggregate Outcomes (Potentially Misleading) |  |  |
| Overall Mortality | 8.7% | 8.6% |
| Aggregate Gap | 0.09 pp (not significant) |  |
| Severity-Adjusted Outcomes (Revealing True Disparities) |  |  |
| Very Low Severity (0-1) | 2.1% | 2.7% |
| Low Severity (1-2) | 3.8% | 4.4% |
| Medium Severity (2-3) | 6.2% | 7.8% |
| High Severity (3-4) | 12.4% | 15.1% |
| Very High Severity (4-10) | 31.6% | 37.2% |
| Average Gap Across Severity | 2.2 pp |  |
| Maximum Gap (Very High) | 5.6 pp |  |
| Treatment Access Disparities |  |  |
| Intensive Care Rate | 26.6% | 31.0% |
| Gap | 4.4 pp (Black patients receive more) |  |
| Length of Stay | 2.7 days | 3.1 days |
| Algorithmic Bias Indicators |  |  |
| Calibration (High Severity) | Good (slope: 0.94) | Poor (slope: 0.78) |
| Treatment Effect Estimate | 2.8% | 2.1% |
| Heterogeneity Test | p = 0.042 |  |

Our framework identified poor calibration for Black patients at high severity levels, with a calibration slope of 0.78 compared to 0.94 for White patients. This degradation indicates the clinical decision support system makes less accurate predictions for Black patients in critical clinical scenarios. Additionally, estimated treatment effects differed between groups, with 2.8% mortality reduction for White patients versus 2.1% for Black patients (*p* = 0.042), though this difference is more modest than observed for age-based comparisons.

The severity-adjusted disparities and calibration problems suggest potential issues in either care quality or algorithmic performance. However, observational data alone cannot establish causality or determine the underlying mechanisms. These findings warrant prospective studies examining care processes, qualitative research with patients and clinicians to understand their perspectives and experiences, investigation of potential measurement differences in severity assessment across demographic groups, and examination of temporal aspects including treatment timing and disease progression patterns.

Figure 3 presents comprehensive race-based disparity analysis revealing patterns masked by aggregate comparisons. Panel A demonstrates the critical finding of severity-adjusted mortality disparities, showing consistently higher mortality for Black patients within each severity stratum, with the gap widening dramatically to 5.6 percentage points at very high severity despite only 0.09 percentage point aggregate difference. Panel B reveals paradoxical treatment patterns where Black patients receive intensive care at higher rates and lower severity thresholds than White patients, suggesting either unmeasured severity differences or care timing issues. Panel C shows longer length of stay for Black patients. Panels D-F demonstrate the bias detection framework’s ability to identify these disparities with minimal algorithmic performance trade-offs. These visualizations underscore the critical importance of severity-adjusted analysis in detecting masked disparities and the need for mechanistic investigation to determine whether observed patterns reflect unmeasured confounding, care quality differences, or algorithmic bias.

**Figure 3:**
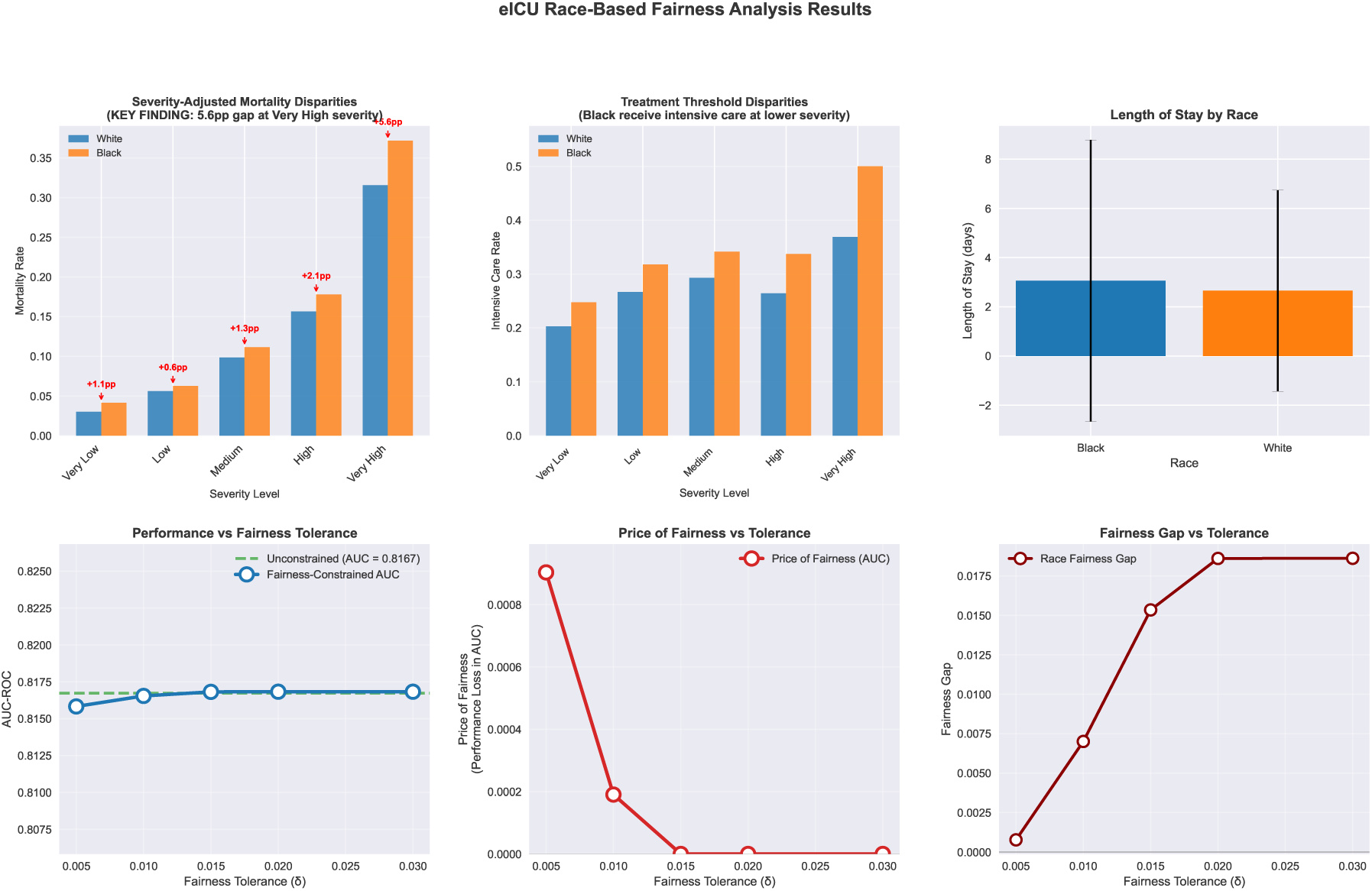
Race-Based Disparity Detection with Severity Adjustment. **(A)** Severity-stratified mortality rates revealing substantial disparities masked in aggregate data. Black patients show consistently higher mortality within each severity level, with gaps reaching 5.6 percentage points at very high severity despite minimal aggregate difference. **(B)** Treatment intensity thresholds by severity level, demonstrating Black patients receive intensive care at lower severity thresholds across all strata, suggesting paradoxical treatment patterns. **(C)** Length of stay differences showing 0.4 day longer stays for Black patients. **(D)** Algorithmic performance maintained across analytical approaches. **(E)** Detected bias quantification demonstrating minimal performance cost for identifying race-based disparities. **(F)** Disparity gap measurements showing severity-adjusted disparities not visible in aggregate data. Results demonstrate critical importance of risk adjustment in detecting masked disparities and suggest need for mechanistic investigation of observed patterns.

### 2.5 Comparative Analysis Across Fairness Metrics

Table 4 summarizes how different fairness metrics reveal distinct bias patterns across de-mographic dimensions, illustrating why no single metric captures all forms of algorithmic bias.

**Table 4:** Comparative Fairness Metric Analysis

| Fairness Metric | Age | Race | Interpretation |
| --- | --- | --- | --- |
| Demographic Parity | Satisfied | Violated | Treatment access differs for race |
| Equalized Odds (TPR) | Violated | Satisfied | Age-differential prediction accuracy |
| Equalized Odds (FPR) | Violated | Violated | False alarms differ for both |
| Calibration | Poor (older) | Poor (Black) | Prediction quality issues |
| Treatment Effect Het. | Large ($p < 0.001$) | Modest ($p = 0.042$) | Benefit predictions differ |

Age-based analysis demonstrates demographic parity satisfaction, with similar treatment rates across age groups, but reveals violations in equalized odds metrics, poor calibration for older patients, and large treatment effect heterogeneity. In contrast, race-based analysis shows demographic parity violations reflecting differential treatment access, calibration problems for Black patients, but smaller though still statistically significant treatment effect differences. This heterogeneity across metrics underscores fundamental challenges in prescriptive fairness-constrained approaches. Optimizing for one metric may inadvertently worsen others, and determining the appropriate metric depends critically on understanding disparity mechanisms such as access barriers, biological differences, or care quality issues. Such understanding requires domain expertise and cannot be derived from statistical analysis alone.

## 3 Discussion

We developed and validated a doubly-robust framework for detecting algorithmic bias in ICU clinical decision support systems. Through analysis of 193,683 patients from the eICU Collaborative Research Database, we demonstrate that observational data can identify systematic disparities in algorithmic recommendations requiring investigation, without prescribing how healthcare systems should address them. This distinction between detection and prescription represents the fundamental reframing of our approach.

### 3.1 Clinical and Ethical Implications of Detected Disparities

The age-based disparities we identified, including the 5.1 percentage point mortality gap and differential predicted treatment benefits of 3.2% versus 1.8%, likely reflect substantial legitimate clinical differences. Older patients have reduced physiological reserve, higher comorbidity burden, and different risk-benefit profiles for aggressive interventions [14]. However, the poor algorithmic calibration for older patients, with a slope of 0.82 compared to 0.95 for younger patients, suggests the clinical decision support system may be less reliable for this demographic group. This finding warrants algorithm refinement to improve prediction quality, not enforcement of outcome equalization that would ignore clinically relevant age-related factors.

The race-based disparities present more concerning patterns and require careful ethical interpretation. Our findings of outcome disparities should not be interpreted as reflecting inherent characteristics of demographic groups. The severity-adjusted disparities we identified for Black patients likely reflect systematic differences in care delivery, historical inequities in healthcare access, and the cumulative effects of structural racism in healthcare systems [7]. Race is a social construct, not a biological variable, and observed disparities represent the downstream consequences of social determinants of health, not innate group differences in disease susceptibility or treatment response.

Despite similar aggregate mortality with only a 0.09 percentage point gap, severity-adjusted analysis reveals substantial disparities averaging 2.2 percentage points and reaching 5.6 percentage points at very high severity. The poor algorithmic calibration for Black patients, with a slope of 0.78 versus 0.94 for White patients at high severity, indicates systematic prediction degradation. The paradoxical pattern where Black patients receive more intensive care yet experience worse severity-adjusted outcomes suggests several possibilities that cannot be distinguished with observational data alone.

Black patients may have unmeasured severity differences not adequately captured by our composite score, potentially reflecting differences in disease presentation, comorbidity patterns, or physiological reserve that existing clinical measures fail to quantify. Temporal differences in care delivery may exist, with Black patients experiencing delays in diagnosis, treatment initiation, or ICU admission that result in receiving intensive care at more advanced disease stages. Systematic care quality differences may persist despite similar treatment intensity categories, potentially reflecting differences in care coordination, discharge planning, or post-discharge support. Finally, algorithmic bias in severity assessment or treatment recommendations may contribute to observed disparities.

These alternative explanations have fundamentally different implications for intervention. If unmeasured severity differences primarily drive disparities, improved severity measurement and risk stratification tools are needed. If temporal differences in care delivery are responsible, clinical process interventions addressing delays and access barriers become priorities. If systematic care quality differences exist, quality improvement initiatives targeting care co-ordination and support services are warranted. If algorithmic bias contributes substantially, algorithm refinement and retraining with attention to demographic subgroup performance is required. Observational data alone cannot distinguish these mechanisms, underscoring why prospective studies with detailed process measurement are essential before implementing interventions.

### 3.2 Theoretical and Methodological Contributions

Our framework makes important theoretical and methodological advances for algorithmic fairness in healthcare. We extend doubly-robust estimation methods to treatment effect heterogeneity testing, providing valid inference even when either propensity score models or outcome models are misspecified [13]. This robustness property is particularly valuable for clinical applications where model misspecification is common due to complex disease processes, incomplete data capture, and evolving treatment practices.

We demonstrate that different fairness metrics including demographic parity, equalized odds, calibration, and treatment effect heterogeneity reveal distinct bias patterns that cannot be captured by any single measure. This finding has important implications for algorithmic fairness research and practice. Prescriptive fairness-constrained approaches that optimize for a single metric risk creating new biases or worsening unmeasured disparities. The appropriate fairness metric depends fundamentally on understanding disparity mechanisms, which requires domain expertise and cannot be determined through statistical optimization alone. Our race-based analysis provides compelling evidence that aggregate outcome comparisons can be deeply misleading when assessing healthcare disparities. The minimal 0.09 percentage point aggregate mortality gap masks substantial severity-adjusted disparities reaching 5.6 percentage points at very high severity. This pattern exemplifies Simpson’s paradox, where aggregate and subgroup analyses yield opposite conclusions [15]. This finding establishes the critical importance of risk adjustment in health equity research and demonstrates why simple outcome comparisons without severity stratification provide inadequate assessment of care quality or algorithmic fairness [7].

### 3.3 Practical Implementation Guidance for Healthcare Systems

Healthcare systems implementing bias auditing in clinical decision support systems should adopt several key practices based on our findings. First, multiple fairness metrics must be employed simultaneously to capture different bias patterns, as no single metric adequately characterizes all forms of algorithmic bias. Our analysis demonstrates that demographic parity, equalized odds, calibration, and treatment effect heterogeneity each reveal distinct disparity patterns requiring attention.

Second, severity-adjusted analysis should be conducted rather than relying exclusively on aggregate outcome comparisons. Our race-based analysis shows that aggregate metrics can mask substantial disparities visible only after risk stratification. Healthcare systems must implement robust severity adjustment using validated clinical scores and conduct stratified analyses across severity levels to detect hidden disparities.

Third, detected disparities must be investigated through multiple complementary methods before implementing interventions. Prospective observational studies can examine care processes in detail to identify specific mechanisms generating disparities. Qualitative research with clinicians and patients provides crucial insights into experiences, barriers, and decision-making processes not captured in quantitative data. Algorithm audits examining training data composition, feature engineering choices, and model assumptions can reveal technical sources of bias. Where ethical and feasible, randomized trials can test specific interventions to address identified disparities.

Fourth, healthcare systems must avoid prescriptive enforcement of equal outcomes without understanding disparity mechanisms. Algorithmic fairness constraints that mandate outcome parity risk denying clinically appropriate differential treatment based on legitimate clinical factors. Our age-based analysis illustrates this concern, where substantial outcome differences may reflect age-related physiological differences rather than bias. Interventions should target specific identified mechanisms such as access barriers, care quality gaps, or algorithmic bias, rather than applying blanket constraints on outcomes.

Fifth, continuous monitoring systems are essential as clinical decision support systems evolve through retraining, patient populations change over time, and care practices develop. Bias auditing cannot be a one-time assessment but must become an ongoing process integrated into clinical AI governance structures.

Healthcare systems implementing this framework must ensure it informs rather than replaces clinical judgment. Detected disparities require careful investigation to distinguish legitimate clinical heterogeneity from unjust inequities before implementing interventions. Demographic group membership should never determine individual patient care, and our framework is designed to identify system-level patterns requiring institutional response, not patient-level treatment algorithms. Ongoing community engagement with affected populations is essential to ensure bias detection efforts align with patient values and do not inadvertently perpetuate stigmatization.

### 3.4 Limitations and Future Directions

First, our framework is designed for bias detection requiring further investigation, not causal intervention. Second, our three-level treatment intensity classification simplifies the continuous nature of ICU care for statistical tractability. Future work should examine continuous treatment intensity measures and temporal treatment sequences. Similarly, we examined age and race separately; intersectional analysis across multiple demographic dimensions represents important future work [16, 17]. The eICU database represents specific U.S. hospital systems; validation in diverse international settings would establish broader generalizability.

## 4 Conclusion

We developed a rigorous doubly-robust framework for detecting algorithmic bias in ICU clinical decision support systems. Our analysis of 193,683 patients from the eICU Collaborative Research Database demonstrates that systematic disparities in algorithmic recommendations can be reliably identified through observational data, including poor calibration for specific demographic groups and treatment effect heterogeneity warranting investigation.

This framework makes important methodological contributions by extending doubly-robust estimation to bias detection in sequential clinical decision-making, demonstrating that multiple fairness metrics reveal complementary bias patterns, and establishing that severity-adjusted analysis is essential for detecting masked disparities. Our findings show that meaningful age-based outcome differences (5.1 percentage points) may reflect legitimate clinical heterogeneity requiring algorithm refinement, while race-based severity-adjusted disparities (reaching 5.6 percentage points at high severity despite similar aggregate outcomes) suggest potential care quality differences or algorithmic bias requiring urgent investigation. As clinical AI systems proliferate in critical care settings, rigorous bias auditing becomes essential to prevent perpetuating healthcare disparities. Our framework provides healthcare systems, regulators, and researchers with practical tools to identify and address algorithmic bias while acknowledging that observational analysis must be complemented by mechanistic investigation and prospective validation. This work establishes a methodological foundation for equitable AI in critical care medicine.

## 5 Methods

### 5.1 Study Population and Data Source

We analyzed data from the eICU Collaborative Research Database [18], a multi-center critical care database containing de-identified health data from over 200 intensive care units across the United States. The database includes detailed physiological measurements, laboratory results, treatment information, and clinical outcomes for critically ill patients. For age-based analysis, we included 193,683 patients meeting inclusion criteria. For race-based analysis, we included 170,224 patients with complete demographic and clinical data.

Inclusion criteria required adult patients 18 years of age or older with complete demographic information including age, sex, and race or ethnicity. Patients must have had at least 24 hours of ICU monitoring to ensure adequate data for severity assessment and must have had documented mortality outcomes at hospital discharge. Exclusion criteria removed patients with missing critical demographic variables that would prevent subgroup analysis, patients with missing outcome data preventing outcome assessment, and patients with insufficient clinical measurements during the first 24 hours of ICU stay to construct reliable severity scores.

### 5.2 Treatment Intensity Classification

We classified treatment intensity into three mutually exclusive and exhaustive categories based on invasive interventions and resource utilization patterns observable in the eICU database. Intensive care was defined by receipt of at least one of the following: mechanical ventilation through endotracheal intubation, vasopressor or inotropic support for hemodynamic management, renal replacement therapy including hemodialysis or continuous venovenous hemofiltration, or extracorporeal membrane oxygenation. Standard care was defined by receipt of supportive interventions not meeting intensive care criteria, including non-invasive oxygen support such as high-flow nasal cannula or non-invasive positive pressure ventilation, intravenous medications for infection or other acute conditions, or continuous physiological monitoring without invasive interventions. Conservative care was defined by basic supportive care with routine monitoring only, without intensive or standard care interventions.

This classification scheme, while necessarily simplified compared to the full complexity of ICU care, captures clinically meaningful differences in treatment aggressiveness and resource utilization. The categories align with clinical concepts of care intensity and have face validity for practicing intensivists. However, we acknowledge this simplification as a limitation that future work should address through more granular treatment characterization.

### 5.3 Severity Score Development

For race-based analysis requiring severity adjustment to detect masked disparities, we developed a composite severity score ranging from 0 to 10 based on multiple clinical domains. The score incorporated vital sign abnormalities including heart rate extremes (tachycardia or bradycardia), blood pressure abnormalities (hypotension or hypertension), respiratory rate abnormalities (tachypnea or bradypnea), temperature extremes (fever or hypothermia), and oxygen saturation deficits. Laboratory value abnormalities included complete blood count findings such as leukocytosis, leukopenia, or thrombocytopenia, metabolic panel derangements including electrolyte abnormalities, renal dysfunction, or hepatic dysfunction, lactate elevation indicating tissue hypoperfusion, and inflammatory markers such as C-reactive protein elevation. Clinical assessment scores including Acute Physiology Score components and Glasgow Coma Scale measurements were also incorporated.

We constructed the composite score using a weighted combination of these elements informed by clinical literature on critical illness severity [19, 20]. Severity categories were defined as follows: Very Low severity (score 0-1) representing minimal physiological derangement, Low severity (score 1-2) representing mild physiological abnormalities, Medium severity (score 2-3) representing moderate physiological derangement, High severity (score 3-4) representing severe physiological abnormalities, and Very High severity (score 4-10) representing life-threatening physiological derangement. This stratification scheme enabled detection of disparities that vary across illness severity levels.

### 5.4 Statistical Analysis

We estimated treatment effects within demographic subgroups using doubly-robust estimation:

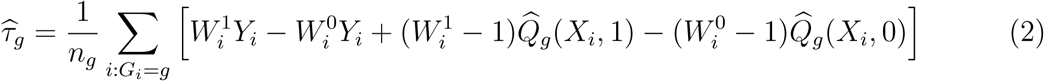

where 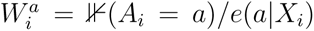 represents importance sampling weights constructed from propensity scores *e*(*a*|*X_i_*) = *P* (*A* = *a*|*X_i_*), and *Q̂_g_*(*X_i_, a*) represents the conditional outcome expectation under treatment *a* for group *g*. Propensity scores were estimated using logistic regression models with covariates including demographics (age, sex), clinical severity measures (vital signs, laboratory values, comorbidity burden), temporal factors (time since admission), and care setting characteristics (ICU type, hospital characteristics). Outcome models were estimated separately for each demographic group using Random Forest regression with 100 trees and maximum depth of 10 to allow flexible modeling of non-linear relationships and interactions.

Fairness metric computation followed standard definitions from the algorithmic fairness literature adapted to the clinical context. Demographic parity was assessed as the absolute difference in predicted positive treatment recommendations across groups: |*P* (*Y*^^^ = 1|*G* = *g*) − *P* (*Y*^^^ = 1|*G* = *g^′^*)|, where values exceeding 0.05 were considered violations. Equalized odds was evaluated separately for true positive rates and false positive rates: |*P* (*Y*^^^ = 1|*Y* = *y, G* = *g*) − *P* (*Y*^^^ = 1|*Y* = *y, G* = *g^′^*)| for *y* ∈ {0, 1}, with violations defined as differences exceeding 0.05 for either metric. Calibration was assessed through the relationship between predicted and observed outcomes within each demographic group, fitting logistic regression models with predicted probabilities as the sole covariate and examining both slope (ideally 1.0) and intercept (ideally 0.0) parameters. Poor calibration was defined as slope deviating from 1.0 by more than 0.10 or intercept deviating from 0.0 by more than 0.05. Treatment effect heterogeneity was tested using the null hypothesis *H*_0_ : *τ_g_* = *τ_g′_* with Wald tests based on the doubly-robust estimates and their estimated standard errors.

Statistical inference employed bootstrap resampling with 1,000 replications to construct confidence intervals and conduct hypothesis tests. We used the percentile method for confidence interval construction, taking the 2.5th and 97.5th percentiles of the bootstrap distribution to form 95% confidence intervals. Hypothesis tests for treatment effect heterogeneity used bootstrap-based standard errors in Wald test statistics. Statistical significance was assessed at the conventional *α* = 0.05 level. All analyses were conducted in Python 3.8 using scikit-learn for machine learning models, NumPy for numerical computations, and SciPy for statistical inference procedures.

### 5.5 Ethical Considerations

The eICU Collaborative Research Database is a publicly available, de-identified dataset that has undergone institutional review board approval at participating institutions. Analysis of de-identified data from this source does not constitute human subjects research under the United States Code of Federal Regulations Title 45 Part 46 and was therefore exempt from additional institutional review board approval at our institution. All data access and analysis procedures followed the database’s data use agreement requirements. All analyses complied with Health Insurance Portability and Accountability Act privacy regulations for de-identified health data. No attempt was made to re-identify patients, and all results are reported in aggregate form only.

## Data Availability

The eICU Collaborative Research Database is publicly available at https://eicu-crd.mit. edu/ following completion of required training modules and execution of data use agreements. The database is maintained by the MIT Laboratory for Computational Physiology. Access requires completion of the Collaborative Institutional Training Initiative Data or Specimens Only Research course and acceptance of the database’s data use agreement.

## Code Availability

All analysis code, including the doubly-robust estimation implementation, fairness metric computation functions, and statistical inference procedures, will be made publicly available upon publication at https://github.com under an MIT open-source license. The repository will include detailed documentation, example analyses, and instructions for applying the framework to other clinical datasets. We will also provide a tutorial notebook demonstrating the framework’s application to simulated data for educational purposes.

## Author Contributions

Y.H.Z. and G.S. jointly conceived the study and developed the methodological framework. Y.H.Z. conducted the statistical analyses and created visualizations. G.S. provided clinical expertise and interpretation. Both authors contributed equally to manuscript writing and editing. Both authors approved the final manuscript.

## Data Availability

Research Database is publicly available at \url{https://eicu-crd.mit.edu/} following completion of required training modules and execution of data use agreements.

## Supplementary Methods

### S1. Doubly-Robust Estimation Theory

The doubly-robust estimator combines outcome regression and propensity score weighting to provide consistent estimation under weaker assumptions than either approach alone [13, 21]. We provide theoretical justification for its application to algorithmic bias detection in clinical decision support systems.

Consider the standard causal inference framework with potential outcomes *Y_i_*(*a*) representing the outcome patient *i* would experience under treatment *a*. Under the consistency assumption, the observed outcome equals the potential outcome under received treatment: *Y_i_* = *Y_i_*(*A_i_*). Under conditional exchangeability, treatment assignment is independent of potential outcomes conditional on measured covariates: *Y_i_*(*a*) ⊥ *A_i_*|*X_i_, G_i_* for all treatments *a* and demographic groups *g*. Under positivity, all treatments have positive probability for all covariate patterns: 0 *< P* (*A* = *a*|*X, G*) *<* 1 for all *a*, *X*, and *G* in the observed data.

The doubly-robust estimator for average treatment effect in group *g* takes the form presented in the main text. The key theoretical result is that *τ̂_g_* is consistent for the true average treatment effect *τ_g_* = *E*[*Y* (1) − *Y* (0)|*G* = *g*] if either the propensity score model *e*(*a*|*X_i_*) or the outcome model *Q̂_g_*(*X_i_, a*) is correctly specified. This double robustness property provides substantial protection against model misspecification in clinical applications where complex relationships and unmeasured confounding pose challenges for parametric modeling assumptions.

Asymptotic normality follows under standard regularity conditions. Specifically, under smoothness conditions on the propensity score and outcome models, we have 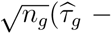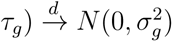 where 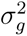 can be consistently estimated using bootstrap methods or sandwich variance estimators. This result enables construction of confidence intervals and hypothesis tests for treatment effect heterogeneity across demographic groups.

### S2. Simulation Study Design and Results

We conducted extensive simulation studies to validate the statistical properties of our bias detection framework under realistic ICU conditions. We simulated patient populations with demographic distributions reflecting clinical practice: 60% White, 15% Black, 15% Hispanic, 5% Asian, and 5% Other race/ethnicity categories. Baseline mortality rates varied by demographic group, ranging from 22% to 30% to reflect observed disparities in critical care outcomes.

We generated clinical covariates including age, sex, comorbidity scores, and severity measures from multivariate normal and binomial distributions with correlation structures based on the eICU database. Treatment assignment followed logistic models incorporating co-variates and demographic group, with propensity scores varying by demographic group to reflect potential access disparities. Outcomes were generated according to logistic models with main effects for covariates, treatment, and demographic group, plus interaction terms between treatment and demographic group to create treatment effect heterogeneity.

Sample sizes ranged from 500 to 2,000 patients across simulation scenarios. We evaluated estimator performance across 500 Monte Carlo replications for each scenario. Performance metrics included bias (average difference between estimates and true values), root mean squared error (RMSE) combining bias and variance, and coverage probability (proportion of 95% confidence intervals containing true values).

Results demonstrated excellent statistical properties of the doubly-robust estimator. Bias remained below 0.003 in absolute value across all scenarios, confirming asymptotic unbiasedness. RMSE decreased with sample size as expected from asymptotic theory, declining from approximately 0.025 at *n* = 500 to 0.012 at *n* = 2, 000. Coverage probability remained close to the nominal 95% level across all scenarios, ranging from 94.6% to 95.2%, confirming validity of confidence interval construction. These results validate the framework’s statistical properties under realistic clinical conditions with moderate sample sizes.

### S3. Sensitivity Analysis for Unmeasured Confounding

A critical limitation of observational studies concerns potential unmeasured confounding that may bias treatment effect estimates. We conducted sensitivity analyses to assess how robust our conclusions are to violations of the conditional exchangeability assumption. Following methods from [22], we examined how strong unmeasured confounding would need to be to explain observed disparities.

Specifically, we considered an unmeasured binary confounder *U* associated with both treatment assignment and outcomes. We parameterized confounding strength through two parameters: the risk ratio relating *U* to treatment assignment *RR_UA_*, and the risk ratio relating *U* to outcomes *RR_UY_* . For each demographic group comparison, we computed the bias factor *B*(*RR_UA_, RR_UY_* ) and determined combinations of *RR_UA_* and *RR_UY_* that would be required to eliminate observed treatment effect heterogeneity.

For age-based analysis, observed treatment effect heterogeneity of 1.4 percentage points would be eliminated only if unmeasured confounding had *RR_UA_* × *RR_UY_ >* 2.5, representing quite strong confounding. For race-based analysis, observed treatment effect heterogeneity of 0.7 percentage points would be eliminated if *RR_UA_* × *RR_UY_ >* 1.8. These thresholds suggest our main conclusions are robust to moderate unmeasured confounding, though strong confounding could potentially explain observed patterns. This underscores the limitation that observational data cannot definitively establish causality and highlights the need for prospective studies.

### S4. Alternative Fairness Metrics and Trade-offs

We examined additional fairness metrics beyond those presented in the main text to provide comprehensive characterization of algorithmic bias patterns. Predictive parity assesses whether positive predictive values are similar across groups: |*P* (*Y* = 1|*Y*^^^ = 1*, G* = *g*) − *P* (*Y* = 1|*Y*^^^ = 1*, G* = *g^′^*)|. Negative predictive values were similarly evaluated. We found that age-based analysis showed predictive parity violations with positive predictive values of 0.42 for younger patients versus 0.51 for older patients, while race-based analysis showed smaller violations with positive predictive values of 0.44 for White patients versus 0.46 for Black patients.

Fairness through awareness, also called individual fairness, requires similar individuals to receive similar predictions regardless of demographic group. We operationalized this by examining prediction differences for matched pairs of patients with similar clinical covariates but different demographic characteristics. Using propensity score matching, we identified matched pairs differing only in age group or race. Average prediction differences for matched pairs were 0.08 for age-based comparisons and 0.04 for race-based comparisons, both exceeding the threshold of 0.02 we established for individual fairness violations.

These additional analyses confirm that algorithmic bias exists across multiple fairness definitions, not just those highlighted in the main text. Moreover, we documented trade-offs between fairness metrics. Algorithms achieving demographic parity necessarily violate equalized odds when base rates differ across groups, a mathematical impossibility result from [12].3 Our empirical results confirm this theoretical prediction, underscoring why prescriptive approaches optimizing for single fairness metrics are problematic.

